# The role of masks, testing and contact tracing in preventing COVID-19 resurgences: a case study from New South Wales, Australia

**DOI:** 10.1101/2020.10.09.20209429

**Authors:** RM Stuart, Romesh G. Abeysuriya, Cliff C. Kerr, Dina Mistry, Daniel J. Klein, Richard Gray, Margaret Hellard, Nick Scott

**Affiliations:** Department of Mathematical Sciences, University of Copenhagen, Copenhagen, Denmark; Disease Elimination Program, Burnet Institute, Melbourne, Victoria, Australia; Institute for Disease Modeling, Global Health Division, Bill & Melinda Gates Foundation, Seattle, USA; School of Physics, University of Sydney, Sydney, New South Wales, Australia; The Kirby Institute, UNSW Sydney, Sydney, New South Wales, Australia; School of Public Health and Preventive Medicine, Monash University, Melbourne, Victoria, Australia; University of Melbourne, Parkville, Victoria, Australia; Department of Infectious Diseases, The Alfred Hospital and Monash University, Melbourne, Victoria, Australia; The School of Population and Global Health and the Peter Doherty Institute for Infection and Immunity, Melbourne, Victoria, Australia

**Keywords:** COVID-19, coronavirus, SARS-CoV-2, modelling, masks, testing, contact tracing, Australia

## Abstract

**Objectives:** The early stages of the COVID-19 pandemic illustrated that SARS-CoV-2, the virus that causes the disease, has the potential to spread exponentially. Therefore, as long as a substantial proportion of the population remains susceptible to infection, the potential for new epidemic waves persists even in settings with low numbers of active COVID-19 infections, unless sufficient countermeasures are in place. We aim to quantify vulnerability to resurgences in COVID-19 transmission under variations in the levels of testing, tracing, and mask usage.

**Setting:** The Australian state of New South Wales, a setting with prolonged low transmission, high mobility, non-universal mask usage, and a well-functioning test-and-trace system.

**Participants:** None (simulation study)

**Results:** We find that the relative impact of masks is greatest when testing and tracing rates are lower (and vice versa). Scenarios with very high testing rates (90% of people with symptoms, plus 90% of people with a known history of contact with a confirmed case) were estimated to lead to a robustly controlled epidemic, with a median of ∼180 infections in total over October 1 – December 31 under high mask uptake scenarios, or 260–1,200 without masks, depending on the efficacy of community contact tracing. However, across comparable levels of mask uptake and contact tracing, the number of infections over this period were projected to be 2-3 times higher if the testing rate was 80% instead of 90%, 8-12 times higher if the testing rate was 65%, or 30-50 times higher with a 50% testing rate. In reality, NSW diagnosed 254 locally-acquired cases over this period, an outcome that had a low probability in the model (4-7%) under the best-case scenarios of extremely high testing (90%), near-perfect community contact tracing (75-100%), and high mask usage (50-75%), but a far higher probability if any of these were at lower levels.

**Conclusions:** Our work suggests that testing, tracing and masks can all be effective means of controlling transmission. A multifaceted strategy that combines all three, alongside continued hygiene and distancing protocols, is likely to be the most robust means of controlling transmission of SARS-CoV-2.

**Strengths and limitations of this study:**

- A key methodological strength of this study is the level of detail in the model that we use, which allows us to capture many of the finer details of the extent to which controlling COVID-19 transmission relies on the balance between testing, contact tracing, and mask usage.
- Another key strength is that our model is stochastic, so we are able to quantify the probability of different epidemiological outcomes under different policy settings.
- A key limitation is the shortage of publicly-available data on the efficacy of contact tracing programs, including data on how many people were contacted for each confirmed index case of COVID-19.

## Introduction

Across the world, governmental responses to the outbreak of the COVID-19 pandemic in the first half of 2020 profoundly curtailed the spread of SARS-CoV-2, the virus that causes the disease (1–4). By midway through the year, an increasing number of countries had moved from an initial crisis-management phase into a new phase centred around minimising transmission risk while allowing societal and economic activities to resume (5). However, by the end of 2020, many countries around the world had experienced epidemic resurgences necessitating further shutdowns (6). This was true even in settings that had come close to eliminating the virus, such as Vietnam (7), New Zealand (8), and Australia (9). In low- or zero-transmission contexts, new outbreaks can emerge if community transmission has not been eliminated, or if infected people arrive from abroad or interstate and interact with the local community (10). It is therefore essential to be able to quantify the risk of epidemic resurgence under different policy settings.

Given the complexities of COVID-19 transmission, including the duration of pre-symptomatic infection (11,12), the proportion of infections that are asymptomatic (13), and the possibility of transmission via surface contact (14), maintaining control of COVID-19 has proven challenging in many jurisdictions (15–19). The often-cited success stories of Taiwan, Vietnam, Thailand, and South Korea included high mask usage, high rates of testing, and fast, effective contact tracing (14–16). The benefits of a multi-pronged approach have also been illustrated in the literature; in the UK, for example, a recent study (23) found that mandating masks in secondary schools could achieve approximately the same reduction in resurgence risk as having an 8-11% increase in symptomatic testing.

In this work, we use an agent-based model to estimate the combination of testing, community-based contact tracing, and mask usage required to maintain epidemic control in a low-transmission, high-mobility setting. These three non-pharmaceutical interventions (NPIs) were key components in reducing the probability of epidemic resurgences prior to the availability of a vaccine, and are likely to remain so for some time even after vaccination coverage increases (24). When used in combination with physical distancing and hand-washing/hygiene measures, all three strategies allow relatively high mobility: testing and contact tracing means that only those at greatest risk of transmitting the virus need to stay home and have been shown to be effective in numerous settings (15,25–29), while masks mean that people with undiagnosed infections present less of a risk to others (30–34).

The context for our study is the Australian state of New South Wales (NSW), with a population of 7.5 million and a cumulative total of just over 4700 diagnosed cases as of December 31, 2020. After an initial wave of COVID-19 infections in March and subsequent lockdown in April, New South Wales began relaxing physical lockdown measures over May and was experiencing near-zero case counts by the start of June, with students back at school, businesses reopening and social/community activities resuming. In late June several clusters of new infections were detected, which subsequently led to a 4-month long period of low but steady case counts, with a mean of ∼5 cases per day over July to October (excluding cases in quarantined travellers). However, NSW subsequently went on to record ∼180 cases in the last two weeks of 2020, a result of a localised outbreak whose containment necessitated the introduction of stringent new restrictions on travel and gatherings that affected many over the holiday period (35).

During the prolonged period of low transmission that NSW experienced in the second half of 2020 prior to December 15, 2020, mobility remained high (36) and transmission was controlled via NPIs. Masks were recommended by the government for the general public and made mandatory for staff in various businesses including supermarkets, but were not universally adopted. In a survey undertaken in mid-September, 78% of the NSW population reported wearing a mask at some point in the previous week (37), although CCTV footage from August registered ∼30% of passengers on urban public transport wearing masks (38). At the same time, high levels of testing were in place, with ∼20,000 people tested per day over June–September (∼2.7/day per 1,000 people), resulting in an average testing yield of 0.05%, one of the world’s lowest (39). The state also had a strong focus on contact tracing, with all cases interviewed within one day of case notification (40). Over the four months from June 1 – September 30, 2020, ∼900 new cases were identified, but only 45 (5%) of were classified as “source unknown”, meaning that they were neither acquired overseas/interstate nor linked to known clusters (41).

Relative to many other contact tracing programs across the world, the NSW contact tracing program was differentiated by its extensive efforts to identify a person’s community contacts in addition to their household, social, school, and workplace contacts (42). The state’s health department (NSW Health) required all businesses to have a COVID-19 Safety Plan, and for the majority of public-facing businesses this included a requirement for customers to register their details upon entry. Upon identifying a new case, NSW Health’s contact tracers would then (a) conduct an extended interview to determine all possible venues in which transmission may have occurred, (b) place details of those venues on their website, on social media, and in newspapers, urging people who had been at the venue to self-isolate for 14 days, (c) attempt to contact all people who were registered to have been at venues within a given window of the time that the diagnosed case was known to have been there and instruct them to self-isolate for 14 days (42).

The dynamics of COVID-19 transmission are complex, and in low-transmission settings the probability of maintaining epidemic control depends on numerous factors outside of policy control, including the characteristics of people who get infected: the size of their households, the type of work that they do, and a number of other socio-economic factors that may influence their contact networks, access to testing and capacity to self-isolate. Several studies have pointed to the role of superspreading events and overdispersion of infections in COVID-19 transmission (25,26,43,44). As a result, even with physical distancing, high levels of testing, and rapid contact tracing, there is still a non-zero probability that a sustained outbreak could occur depending on who gets infected and where. In this study, we consider a range of testing and contact tracing levels, and assess the roles of masks, testing and contact tracing as a means of controlling community-based transmission.

## Methods

### Transmission model

We used an open-source agent-based model, Covasim (45), developed by the Institute for Disease Modeling with source code and documentation available at https://covasim.org, and previously adapted by our group to model the Victorian epidemic (10,46). Covasim contains detailed descriptions of age-dependent disease acquisition and progression probabilities, duration of disease by acuity, and the effects of interventions including symptomatic and asymptomatic testing, isolation, contact tracing, and quarantine, as well as other NPIs such as physical distancing, hygiene measures, and protective equipment such as masks. Importantly, it also captures individual variability, with viral loads varying both between individuals and over time.

We began by simulating a population representative of New South Wales by taking data on the age and sex composition of the population from the 2016 census (the latest available), and using it to create a model population of agents with similar characteristics. The simulations consist of 100,000 individual agents, who are dynamically scaled based on prevalence to represent the total New South Wales population of 7.5 million. The dynamical scaling means that whenever the proportion of susceptible agents falls below a threshold of 5%, the number of agents in the model is increased; further implementation details can be found in Section 2.3.6 of Kerr et al (45).

Next, we created contact networks for these agents. The governmental response to COVID-19 in New South Wales consisted of a set of highly context-specific policies covering individuals, businesses, schools, and other types of organisations. To model these policies, we allow agents in the model to interact over five types of contact network: households, schools, workplaces, and static and dynamic community networks. The static community network consists of interactions with friends, colleagues, or other known associates who come together on a regular and predictable basis, and contains four sub-networks: professional sports, community sports/fitness/leisure clubs, places of worship, and socialising with friends. The dynamic community network consists of interactions in which people interact with strangers or random groups of people, and contains seven separate sub-networks, representing: (1) arts venues such as museums, galleries, theatres, and cinemas, (2) large events such as concerts, festivals, sports games, (3) pubs and bars, (4) cafes and restaurants, (5) public parks and other outdoor settings, (6) public transport, (7) all other community settings. The method for constructing these networks is described in our previous study of the Victorian epidemic (46) and is based on the methodology of the SynthPops Python package (47).

Finally, to simulate the epidemic and policy environment in New South Wales over March 1 – September 30, 2020, we include parameter changes that capture the testing, tracing, isolation, quarantine, and lockdown policies that were enacted over this time. Figure 1 presents a summary of how contact networks and the relative risk of COVID-19 transmission in different settings changed as policies evolved. Some of these changes in transmission risk are derived from available data (42), while others are taken from a similar modelling exercise conducted in Victoria, in which a panel of Australia-based experts reviewed the likely effect of policies on transmission risks (46). Further details of all policies and how we model their effects on transmission risk are contained in Supplementary Table 1. We also model the introduction of interstate cases in late June prior to the closure of the state border with neighbouring Victoria.

**Table 1:**
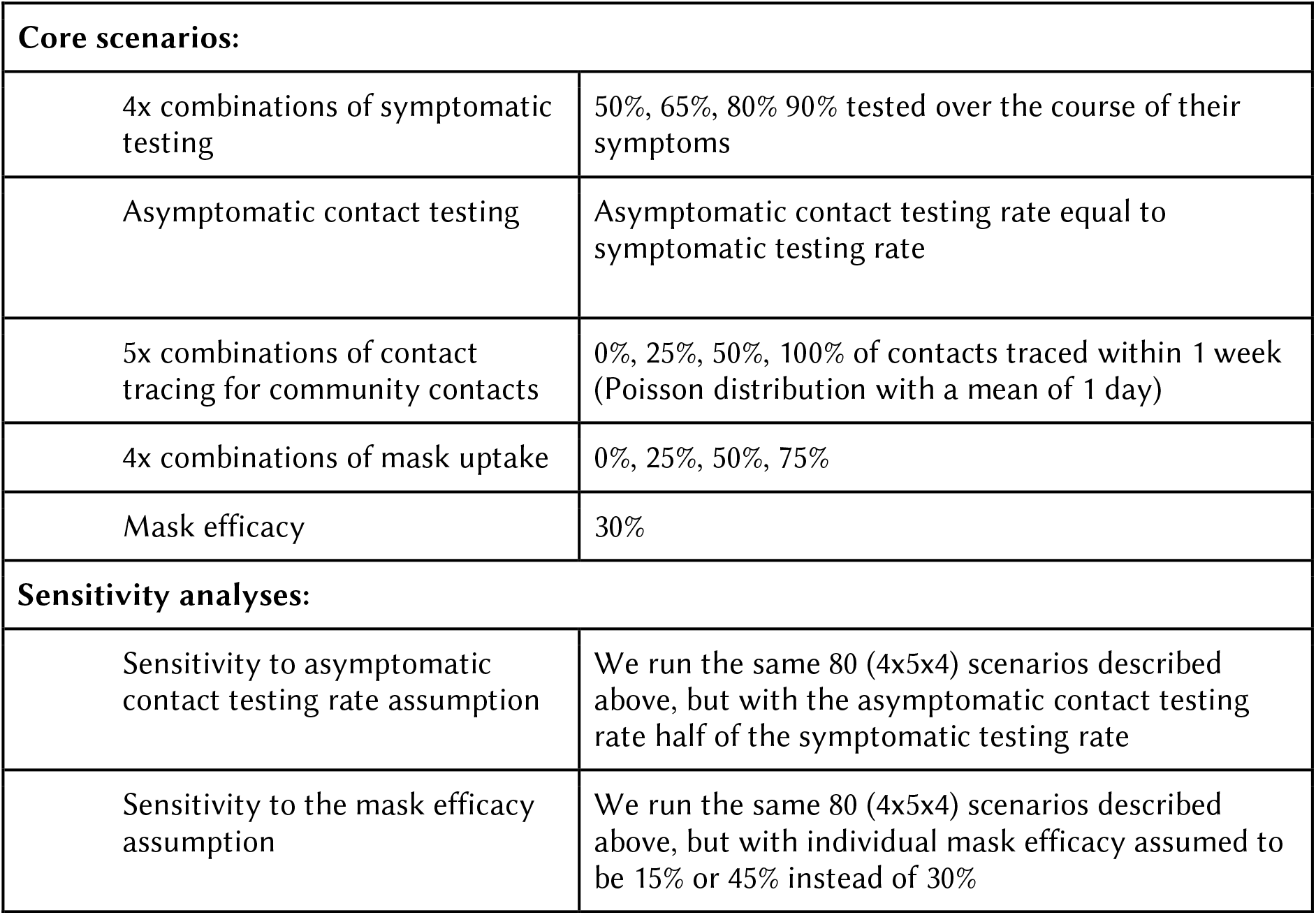
overview of the scenarios analysed over Oct 1 – Dec 31, 2020.

**Figure 1.**
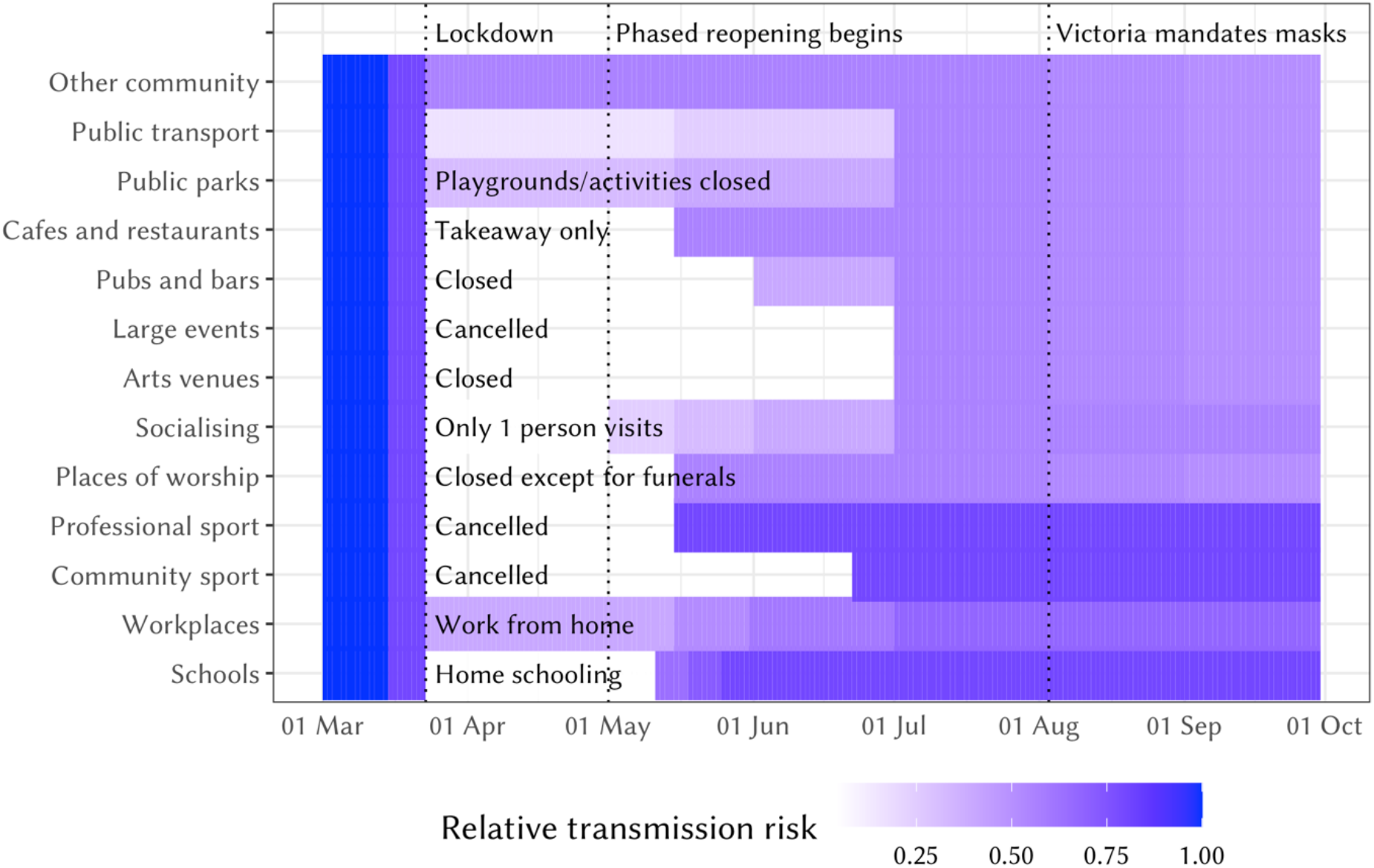
Relative changes in network structure and transmission risk across different settings in New South Wales over March–August. The absolute transmission risk varies by setting and is highest in household and lowest in outdoor settings (see (46) for details).

### Data and model fitting

We calibrate the model by adjusting the per-contact transmission risk to minimise the absolute differences between the model projections and data on the daily number of cases diagnosed, excluding cases acquired overseas or interstate from NSW Health (41). Specifically, we drew 500 samples from a Gaussian prior distribution of values for the per-contact transmission risk (∼N(0.025,0.002)). For each sample, we run N=20 simulations for each to produce 10,000 trial simulations, and then retain the 1% of these with the minimum absolute differences between the model projections and the data.

### Model analysis and timeframe

As described above, we fit the model using data up until September 30, 2020. We then run the model for three months (from October 1, 2020, until December 31, 2020), under a set of assumptions about future testing rates, the efficacy of contact tracing, and mask uptake. For each scenario (described below), we quantify the probability of the epidemic exceeding certain thresholds. We then compare these projections to the observed epidemic outcomes over the period from October 1 to December 31, 2020.

For testing rates, we distinguish between symptom-based testing and testing of asymptomatic contacts. NSW Health guidelines advise anyone with symptoms to get tested, as well as anyone identified as a contact regardless of symptoms (42). To reflect this, we use a baseline assumption that the testing rate for asymptomatic contacts is the same as for people with symptoms, and run a counterfactual set of scenarios in which it is assumed to be half the rate for those with symptoms (Table 1). We modelled scenarios in which 50%, 65%, 80%, or 90% of those with symptoms get tested. Test results are assumed to be communicated within 24 hours (50).

We also considered variations on mask uptake and effectiveness. On uptake, we modelled scenarios in which 0%, 25%, 50%, or 75% of the population will wear masks in dynamic community settings (i.e., settings in which people interact with strangers or random groups of people), which in the model includes arts venues, large events such as concerts, festivals, sports games, public parks and other outdoor settings, public transport, and all other community settings. We do not consider scenarios in which 100% of the population wear masks across these settings, due to the infeasibility of wearing masks whilst eating or drinking. On efficacy, we note that although the body of evidence supporting the effectiveness of masks for protecting against transmission between individuals is now considerable, the size of the effect is difficult to determine, with estimates in the range of 20–80% and varying depending on whether one or both parties wear masks (31,49,51), or whether spillover behavioural changes on people’s attention to other NPIs are captured (52). To capture the uncertainty regarding the effectiveness of masks, we assume that masks will reduce the per-contact probability of transmission by 30%, in line with estimates from (53), but also consider 15% and 45% in a sensitivity analysis presented in the supplementary materials.

To model the efficacy of contact tracing, we use publicly-available data from NSW Health (42). We note that, although the program reports the proportion of known contacts that were reached within defined timeframes, we would ideally like to know the proportion of all contacts that were reached, which will be lower than the reported values since it will also include contacts that the case did not recall or disclose. Thus, although NSW Health’s published reports (42) indicate that 100% of contacts are notified within 48 hours, we use slightly more conservative values, namely that 100% of household contacts will be traced and notified on the same day that test results are communicated, and that 95% of school contacts and 90% of workplace contacts will be notified on the following day. We then consider scenarios in which 0%, 25%, 50%, 75%, or 100% of all other contacts (which we refer to as community contacts) can be traced within a week of a case notification. For each scenario, the time to trace each contact is drawn from a scaled Poisson distribution with a mean of 1 day (Figure S1). We run 100 simulations for each permutation of testing rates, mask usage and contact tracing efficacy.

Finally, we assume that people who have been contact-traced will quarantine with 90% compliance from their workplace, school, and community contacts. Whilst this assumption may be optimistic in other global contexts, the lower case counts in NSW mean that contact tracers have far greater capacity for rigorous ongoing follow-up of contacts, and breaches of isolation are escalated with local authorities, as stipulated in national guideline documents (50).

#### Patient and public involvement

This is a modelling study; no personal data was used so patient/public involvement was not required.

## Results

According to our modelled estimates, 4,550 (95% projected interval: 3,700–6,660) people had acquired COVID-19 in NSW by the end of September 2020 (excluding cases acquired overseas or interstate), of which 39% (27–48%) had been diagnosed (Figure 2). The majority of undiagnosed infections occurred during the March-April wave; since the beginning of June, we estimate that 65% of all locally-acquired infections have been diagnosed, with the remaining 35% being primarily comprised of asymptomatic infections (65%). We further estimate that 79,330 (60,100–129,020) people had been required to self-isolate at some point by September 30, 2020 as a result of having potentially been in contact with a confirmed case.

**Figure 2.**
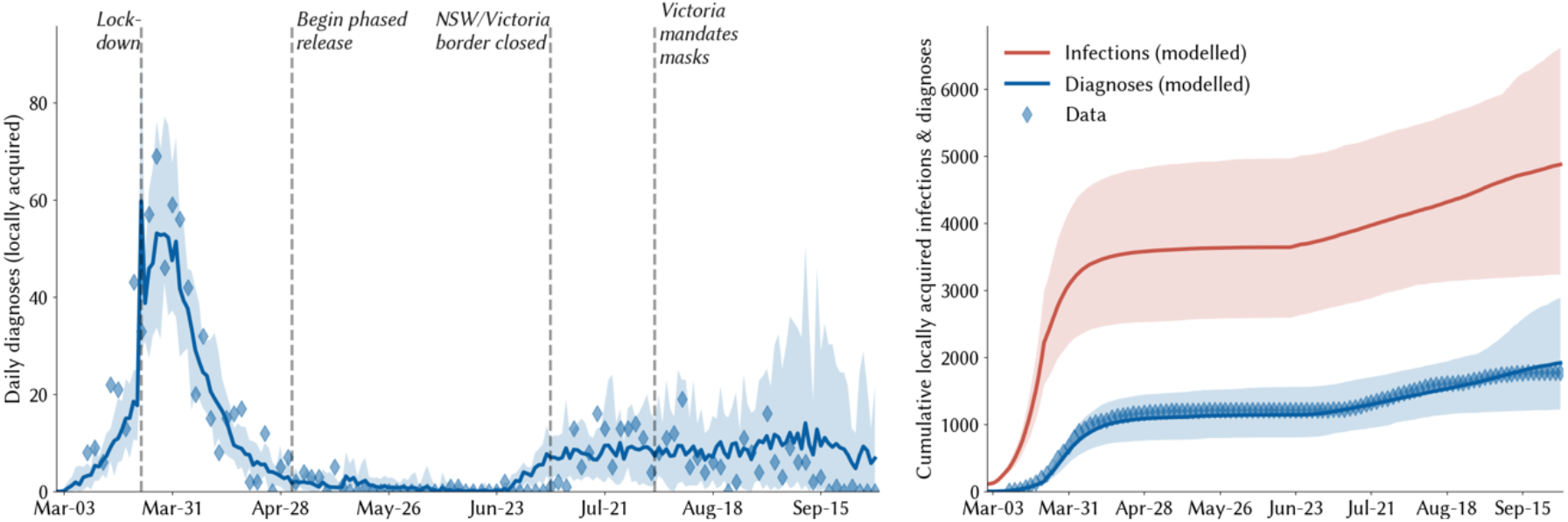
Calibration of the model to the NSW epidemic. Solid lines indicate the median model projections over 100 model runs; shaded areas indicate 95% projected intervals over different initialisations; blue diamonds indicate data on confirmed locally-acquired cases.

Estimates of daily new infections under each combination of testing, community contact tracing, and mask usage are presented in Figure 3. A key finding highlighted by Figure 3 is how effective high levels of testing are in maintaining epidemic control: all strategies in which there is at least some contact tracing in place and testing rates are very high (90% of people with symptoms and 90% of asymptomatic contacts of confirmed cases) lead to a robustly controlled epidemic, with a median of ∼180 infections in total estimated over October 1 – December 31 under high mask uptake scenarios, or 260–1,200 without masks, depending on the efficacy of community contact tracing (Figure 3, bottom row). However, holding mask uptake and contact tracing constant, we estimate that the number of infections over October 1 – December 31, 2020, would be 2-3 times higher if the testing rate was 80% instead of 90%, 8-12 times higher if the testing rate was 65%, or 30-50 times higher with a 50% testing rate (Figure 3, third row).

**Figure 3.**
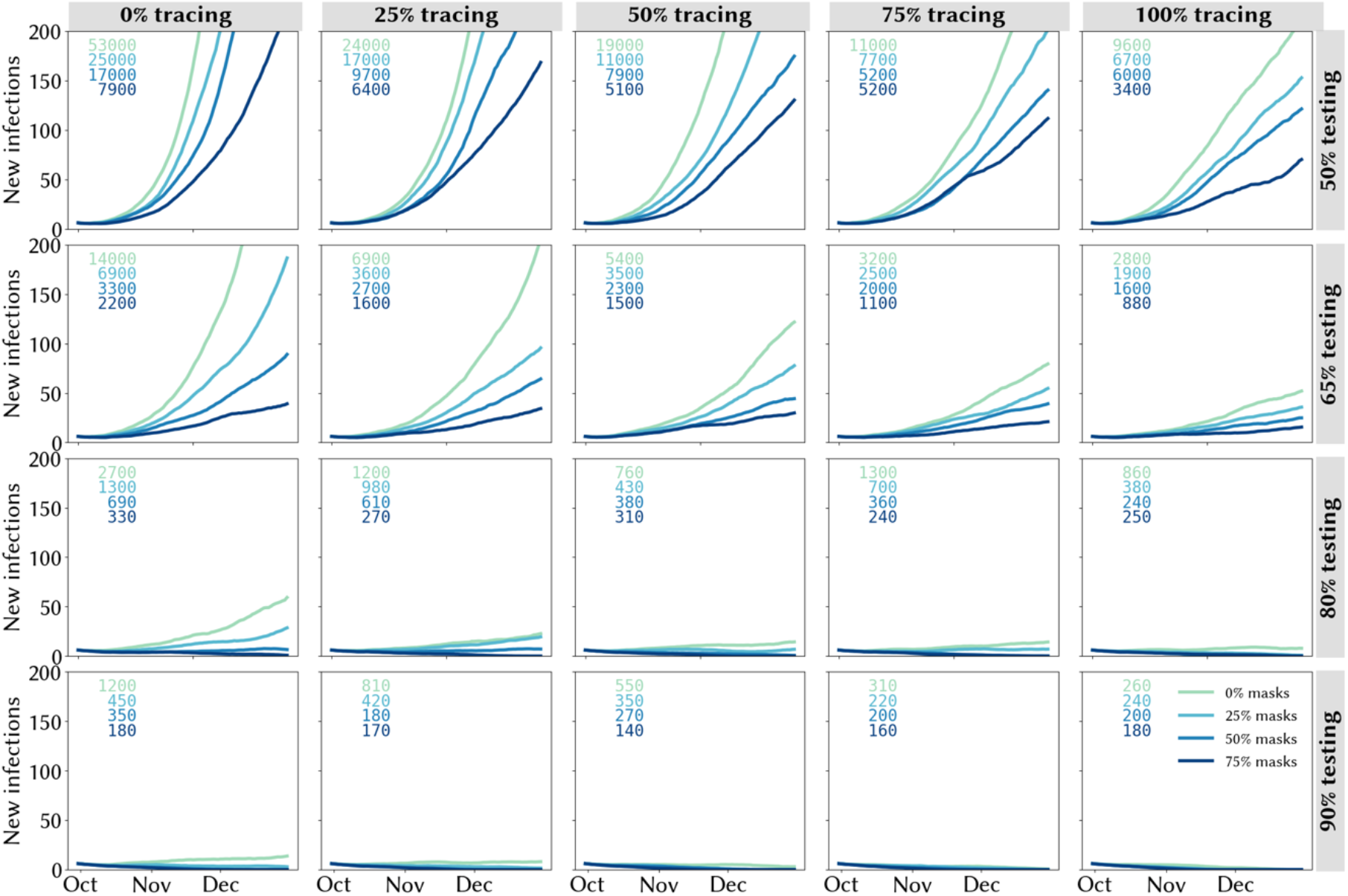
Trailing 14-day average of daily locally-acquired infections under different assumptions about the testing rate (rows), proportion of community contacts that can be traced within one week (columns), and mask uptake (line colours). Projections represent the median of 100 simulations. Text boxes in each panel displays the cumulative number of infections over October 1 – December 31, 2020.

A second key finding is that the lower the testing rate, the greater the impact of masks and contact tracing (and vice versa). With medium-lower testing rates of 50% or 65%, the marginal impact of both masks and contact tracing are considerable (Figure 3, top two rows). Under these scenarios, the most robust strategies consist of a combination of masks and community contact tracing. Assuming a 50% testing rate, a scenario with near-perfect tracing and no masks is approximately equivalent to a scenario with no tracing and high mask uptake. However, without community contact tracing, a reduction in mask usage from 75% to 50% would lead to a near-doubling in estimated infections (from 7,900 to 17,000), whereas even moderate levels of community contact tracing (25% traceable within a week) would increase the robustness of the response to lower mask usage (with 6,400 infections estimated under 75% mask usage and 9,700 under 50% mask usage). Similarly, masks strengthen the resilience of the epidemic outcome to decreases in contact tracing efficacy: without masks, a reduction in community contact tracing from 75% to 50% would lead to a 70% increase in total infections, whereas it would have no impact if 75% of the population were wearing masks.

Figure 4 captures the risks associated with different strategies, quantified in terms of the probability of the 14-day average of daily diagnoses exceeding a given number by December 31, 2020. With no community tracing and a 50% testing rate, the probability of exceeding 100 diagnoses/day by the end of 2020 was calculated to be 98% without masks, and to remain greater than 50% even under the most optimistic mask uptake scenario (Figure 4, top left panel). However, higher levels of community tracing reduce these risks considerably: even without any mask usage, the probability of exceeding 100 cases/day is estimated to be half as high with perfect community tracing (Figure 4, top right panel) compared to the same scenario without community tracing (48% vs 98%), and the addition of mask usage in community settings is estimated to reduce this even further, to ∼10%.

**Figure 4.**
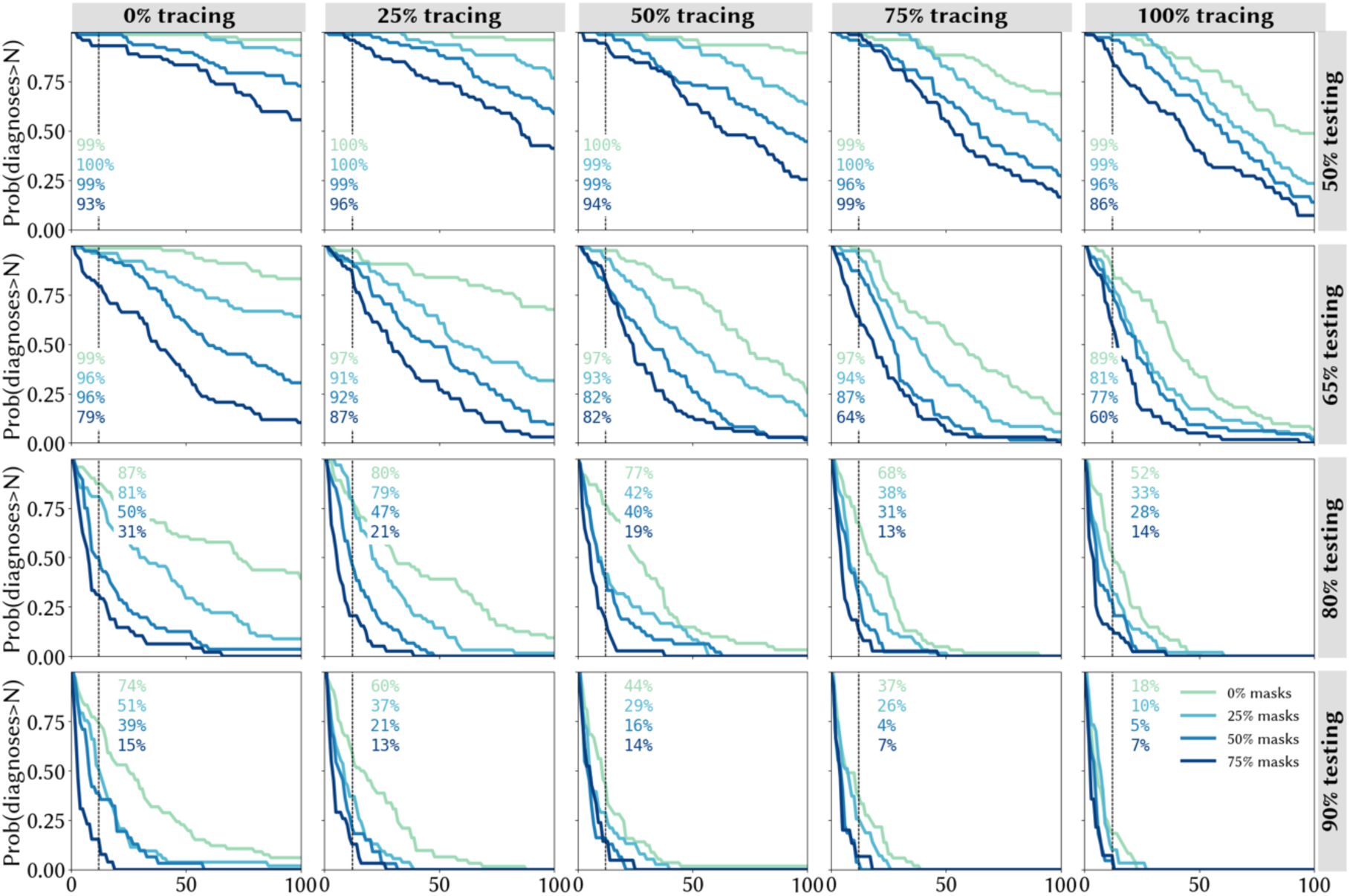
Quantifying the likelihood of epidemic resurgence in New South Wales: the probability of the trailing 14-day average of locally-acquired cases exceeding a given number by December 31, 2020 under different assumptions about the testing rate (rows), proportion of community contacts that can be traced within one week (columns), and mask uptake (line colours).

### Comparison with actual outcomes

The text boxes on Figure 4 display the probability of observing the outcome that was seen in NSW, in which the two-week average of daily diagnoses exceeded 12 by December 31. We calculate that this outcome would have been assigned a low probability (4-7%) under the best-case scenarios of extremely high testing (90%), near-perfect community contact tracing (75-100%), and high mask usage (50-75%), but that it would have been considered a far more likely outcome if any of these were assumed to be at lower levels.

### Sensitivity analyses

If asymptomatic contacts only test at half the rate of people with symptoms, we estimate a more severe epidemic over the last quarter of 2020, but the key results regarding the roles of community contact tracing and masks do not change (Figure S2). Since our core analyses already assume high compliance with recommended self-isolation policies for known contacts of confirmed cases, the marginal benefit of high asymptomatic contact tracing is primarily to further bolster the efficacy of contact tracing, since it allows for the identification of chains of transmission even in the absence of symptoms. To highlight the role of testing asymptomatic contacts, in Figure 5 we highlight a particular set of scenarios corresponding to the most optimistic contact tracing assumptions, in which all community contacts are assumed to be traced within one week, with a mean time to trace of one day (the full set of scenarios are summarised in Figure S2). Within this set of scenarios, the total number of infections is estimated to be around 50% higher if asymptomatic contacts test at a lower rate than people with symptoms (averaged across all levels of mask usage).

**Figure 5.**
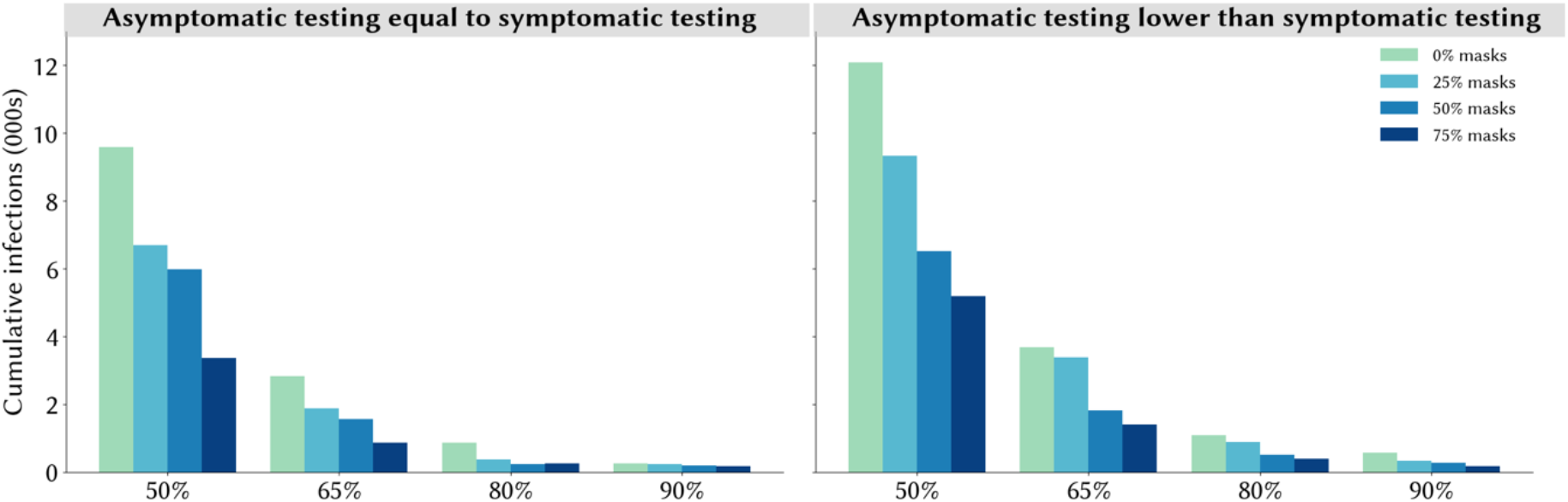
Estimated total infections over October 1 – December 31, 2020 under different assumptions about testing rates and mask uptake, assuming all community contacts can be traced within a week with a mean time to trace of 1 day. Projections represent the median of 100 simulations.

We also find that, if masks are assumed to be more or less effective, this would change the risks associated with diagnosing more than a given number of cases per day under the different mask scenarios, but would not qualitatively change the results (Figure S3-S4).

## Discussion

In this work we presented estimates illustrating the extent to which masks, community testing, and community contact tracing could reduce the spread of SARS-CoV-2 in New South Wales, Australia in the absence of widespread vaccination. We found that the relative impact of masks is greatest when testing and tracing rates are lower (and vice versa). With very high testing rates (90% of people with symptoms, plus 90% of people with a known history of contact with a confirmed case), we estimate that epidemic control would be possible even without widespread mask uptake, provided that fast and effective contact tracing is in place. However, under any scenario where testing rates are lower, we estimate that mask use can play an important role in reducing the potential for epidemic resurgence.

Our findings have relevance for assessing the outbreak that occurred in the state towards the end of December 2020, which saw an average of ∼12 diagnoses/day over two weeks. According to our estimates, this outcome would have been assigned a relatively low probability (4-7%) if it was assumed that compliance with the state’s testing guidelines, venue registration protocols, and non-mandatory mask recommendations would be at very high levels, but that it would have been evaluated as a far more likely outcome if compliance with any of these NPIs was assumed to be lower. As a result of the December 2020 outbreak, the NSW government made masks mandatory in all indoor community settings from January 4, 2021 (54), a decision which will likely increase the robustness of the overall response to decreases in testing or contact tracing rates, but will still require ongoing commitment to ensure these remain at high levels, at least until vaccination coverage is sufficiently high. According to Australia’s vaccination schedule, half the population may be vaccinated by mid-2021, and NPIs will feature as part of the COVID-19 control strategy until well beyond this point (55).

Of the interventions considered, this study suggests that maintaining high levels of symptomatic testing, contact tracing, and testing of contacts are all important. We also quantified the importance of testing asymptomatic contacts as a strategy that further strengthens the power of contact tracing programs, since it allows the tracing and quarantining of their contacts in turn. Longer term, ‘pandemic fatigue’ may bring about challenges in maintaining high levels of testing, mask use, and contact tracing (56). This study suggests that having high levels of any two of mask use, testing and contact tracing can largely mitigate the need for the third; however practical challenges mean that this is unlikely to occur, and a more multifaceted approach of aiming for high coverage of all three and ending up with moderate coverage of all three may be an effective and more robust strategy. Various studies have shown that the roll-out of vaccination plans is unlikely to mitigate the need for NPIs such as these for some time (24,57–59), so these finding are likely to remain relevant for a considerable portion of 2021.

Although various efforts have been made to synthesise the ever-expanding body of research regarding the efficacy of different interventions (1–4,61), each country’s epidemic has distinct characteristics and there are very few standardised, globally-applicable guidelines on what constitutes a best-practice public health strategy (62,63). As a result, jurisdictions have taken diverse approaches in terms of which interventions to prioritise, from a list that includes physical distancing, travel restrictions, wearing of masks or face coverings, isolation and/or testing of those with COVID-like symptoms, isolation of those who test positive, and tracing the contacts of confirmed cases for testing and/or quarantining. Although it may not always be articulated as such, numerous trade-offs are being made between different policy options in an attempt to allow the highest degree of societal activity commensurate with epidemic control. In New South Wales, mask use was encouraged in particular settings since July 2020, but not mandated until January 2021; at the same time, there was a strong focus on contact tracing. The results from this work suggest that the prioritisation of contact tracing may mitigate the relative importance of masks to some extent, but that this relies on continued high levels of community testing.

There are several limitations to this study. Firstly, the mathematical model that we use requires data on various aspects of SARS-CoV-2 transmission and prevention that are still not known exactly, including the effects of masks on preventing individual transmission and the proportion of infections that are asymptomatic. Whilst we have used the best available data and sampled from appropriate distributions where possible, this represents a source of uncertainty in all mathematical models of COVID-19. Additional uncertainties are introduced by the evolution of new strains of SARS-CoV-2 with increased transmissibility and/or severity. Secondly, we have constructed sets of scenarios that examine various combinations of parameters on mask uptake, contact tracing, testing of people with symptoms, and asymptomatic contact testing, but there are many more parameters that determine the dynamics of transmission, including the stringency of border control measures, people’s adherence to quarantine and isolation policies, and the effect of ongoing distancing policies. Changes to any of these policies would affect the results presented here in ways that are not straightforward to predict or extrapolate. Third, we have not considered any outbreak risk associated with newly seeded cases in the community that may arise from international or interstate arrivals. Another limitation is the relatively simplistic way that we have modelled mask usage, whereby we have not included variation in (a) adherence to mask usage across different types of venue, or (b) individual compliance. Assuming that mask-wearing reduces everyone’s transmission risk by a certain percentage disregards the individual behavioural changes that may adjust individual-level transmission risk by varying amounts. Furthermore, it is very likely that mask usage would be higher in certain settings (e.g., public transport) than others, especially restaurants which are suspected to be important for transmission. Finally, the model used here does not contain a geospatial component, and we have not considered heterogeneities in incidence, behaviour, or contact patterns across different parts of the state in these analyses. This could be relevant for questions around mask uptake, as uptake of masks is generally higher in more densely populated areas, so using a state-wide average for the proportion of the population wearing masks may underestimate their impact, especially since >90% of infections in NSW to date have occurred in the Sydney metropolitan area.

## Conclusions

Our work suggests that testing, tracing and masks can be effective means of controlling transmission in dynamic community settings, and higher compliance with one can offset lower compliance with the other to some extent. However, pursuing a strategy that combines aggressive testing, high mask usage, and effective contact tracing, alongside continued hygiene and distancing protocols, is likely to be the most robust means of controlling community-based transmission of SARS-CoV-2.

## Data Availability

All code and data analysed during this study are available in
https://github.com/optimamodel/covid_nsw

## Declarations

### Data sharing statement

All code and data analysed during this study are available in https://github.com/optimamodel/covid_nsw.

### Contributorship

RMS wrote the manuscript and produced the results. The model parameters and calibration were agreed upon by RMS, RA, NS, and RG. The scenarios and analyses were agreed upon by NS, MH, and RMS. CK, RMS, RA, DM, and DJK led development of the model (along with numerous other contributors listed below).

## Acknowledgements

We thank all contributors to the Covasim model, including at the Institute for Disease Modeling, including Brittany Hagedorn, Katherine Rosenfeld, Prashanth Selvaraj, Rafael Nunez, Gregory Hart, Carrie Bennette, Marita Zimmermann, Assaf Oron, Dennis Chao, Michael Famulare, Lauren George; at GitHub, including Michał Jastrzębski, Will Fitzgerald, Cory Gwin, Julian Nadeau, Hamel Husain, Rasmus Wriedt Larsen, Aditya Sharad, and Oege de Moor; at Microsoft, including William Chen, Scott Ayers, and Rolf Harms; and at the Burnet Institute, including Anna Palmer, Dominic Delport, and Sherrie Kelly. This work benefited from a useful discussion with James Wood, Deborah Cromer, and John Kaldor of the University of NSW, Australia.

## Supplementary materials

**Figure S1.**
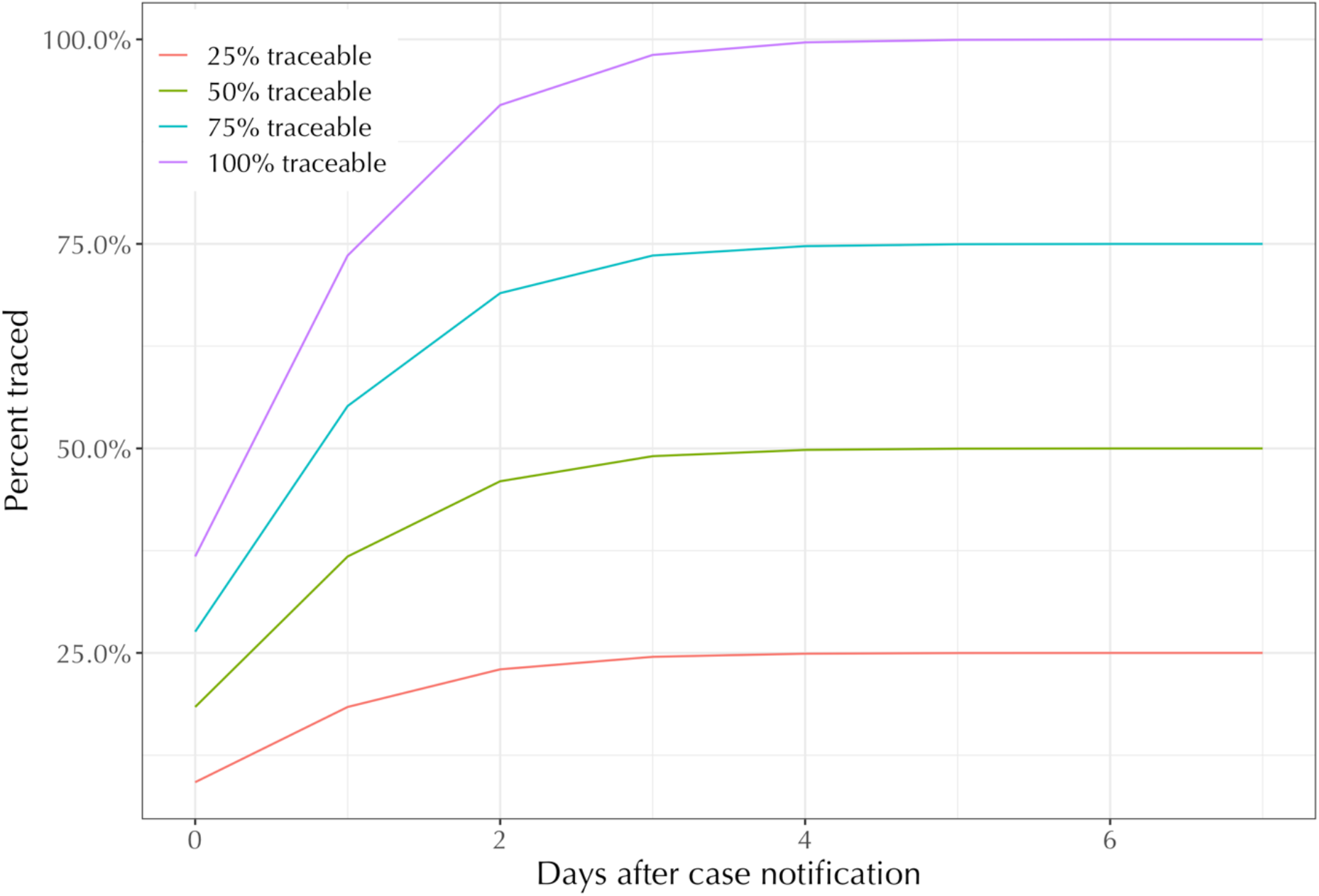
Assumed distribution of times required to find venue-based contacts.

**Table S1.** Effects of policies on transmission risk in New South Wales

| Setting | Description of policy changes and their effects |
| --- | --- |
| Schools | School attendance rates in NSW had already dropped by 25% by 15 March 2020 (1), and on 23 March 2020 the NSW Premier advised that although schools remained open, parents were encouraged to keep their children at home for online learning (2). School attendance rates subsequently dropped to 5% of their pre-COVID levels (3). However, attendance quickly returned to pre-COVID levels shortly after schools reopened in mid-May (4). To model this, we removed 95% of contacts between school children and then restored them again as schools opened, but with the relative transmission risk set to 80% of its pre-March levels to account for additional safety measures in place for school activities (5). |
| Workplaces | According to survey data from the Australian Bureau of Statistics, almost half of working Australians were working from home in late April/early May (6), which is roughly consistent with Google movement data indicating that 40% fewer people were at work over that period compared to baseline. Workplace-based activities increased as COVID restrictions eased, but remained 15% lower over June-July compared to baseline. In the model, we removed 50% of workplace contacts and then restored them so that the workplace network was back to 85% of its pre-COVID size by the beginning of July (Figure 1). As with schools, we set the relative transmission risk set to 80% of its pre-March levels to account for the presence of NPIs. |
| Static community | We assume that almost no contacts occurred over these networks from March 23 to May 1 with the exception of the limited contacts arising from permitted single-person visits. These networks were gradually restored over the period from May 1 to July 9 as restrictions eased. |
| Dynamic community networks over May-July (negligible mask usage) | Within New South Wales, arts venues such as museums, galleries, theatres, and cinemas, large events such as concerts, festivals, sports games, and pubs/bars were all closed over the period from March 23 to May 15, after which the networks were gradually restored. Cafes and restaurants, public parks and other outdoor settings, public transport, and all other community settings including essential retail remained open in some capacity throughout the year but with decreased demand and operational restrictions to reduce the likelihood of transmission (e.g., takeaway service only, closure of playgrounds, capacity limits on transport, and physical distancing/hygiene). |
| Dynamic community networks over August (increased mask usage) | From August 3, 2020, the use of masks was mandated in Victoria, which led to a marked increase in mask usage across the country. Only 13% of Australians wore a mask at least once over the month of June, but 58% reported wearing one at least once over August 7–17, 2020 (99% of Victorian residents compared to 44% of residents of other states) (7). Within New South Wales, media sources reported that 30% of people were wearing face masks on public transport in central Sydney in mid-August (8). To reflect the gradual increase in mask uptake in the model, we adjust the relative transmission risk assuming that the proportion of adults who wore masks while at work and in dynamic community settings increased over August to reach 30% by the end of the month. |
| Testing, tracing, and isolation | We assume that the daily testing probability for those with symptoms increased from 5% in April to 20% by the beginning of June. Assuming a symptomatic period of roughly 10 days, this implies that the proportion of symptomatic people who get tested increased from 40% to 90%. In addition to symptom-based testing, we also assume that 90% of people who are not symptomatic but have been told to quarantine as a result of having been in contact with a confirmed case will get tested. Our assumptions around test sensitivity coupled with our modelling of viral load kinetics are specified such that the probability of identifying a true positive increases and then decreases over the course of an infection, following a similar profile to that reported in the literature (9–10).<br>To model the efficacy of contact tracing over the period from June 1 – September 30, 2020, we assume that all household contacts were traced and notified on the same day that test results were communicated, that 95% of school contacts and 90% of workplace contacts were notified on the following day, and that 50% of all other contacts (which we refer to as community contacts) were traced within a week of a case notification, with a mean time to trace of one day (Figure S1).<br>We assume that confirmed cases will isolate with near-perfect effectiveness, meaning that the probability of them transmitting to school or workplace contacts is zero, and the probability of them transmitting to their household contacts is reduced by 80%. Similarly, we assume high levels of adherence to isolation policies |
|  | imposed on those who have been notified that they were in contact with a confirmed case, with a 90% reduction in their probability of transmitting to school or workplace contacts. |
| Quarantine, and border control measures | From early on in the epidemic, Australia imposed strict border control measures requiring all arrivals to quarantine for two weeks in a hotel. These policies are assumed to be maintained throughout the duration of the model simulations. |

**Figure S2.**
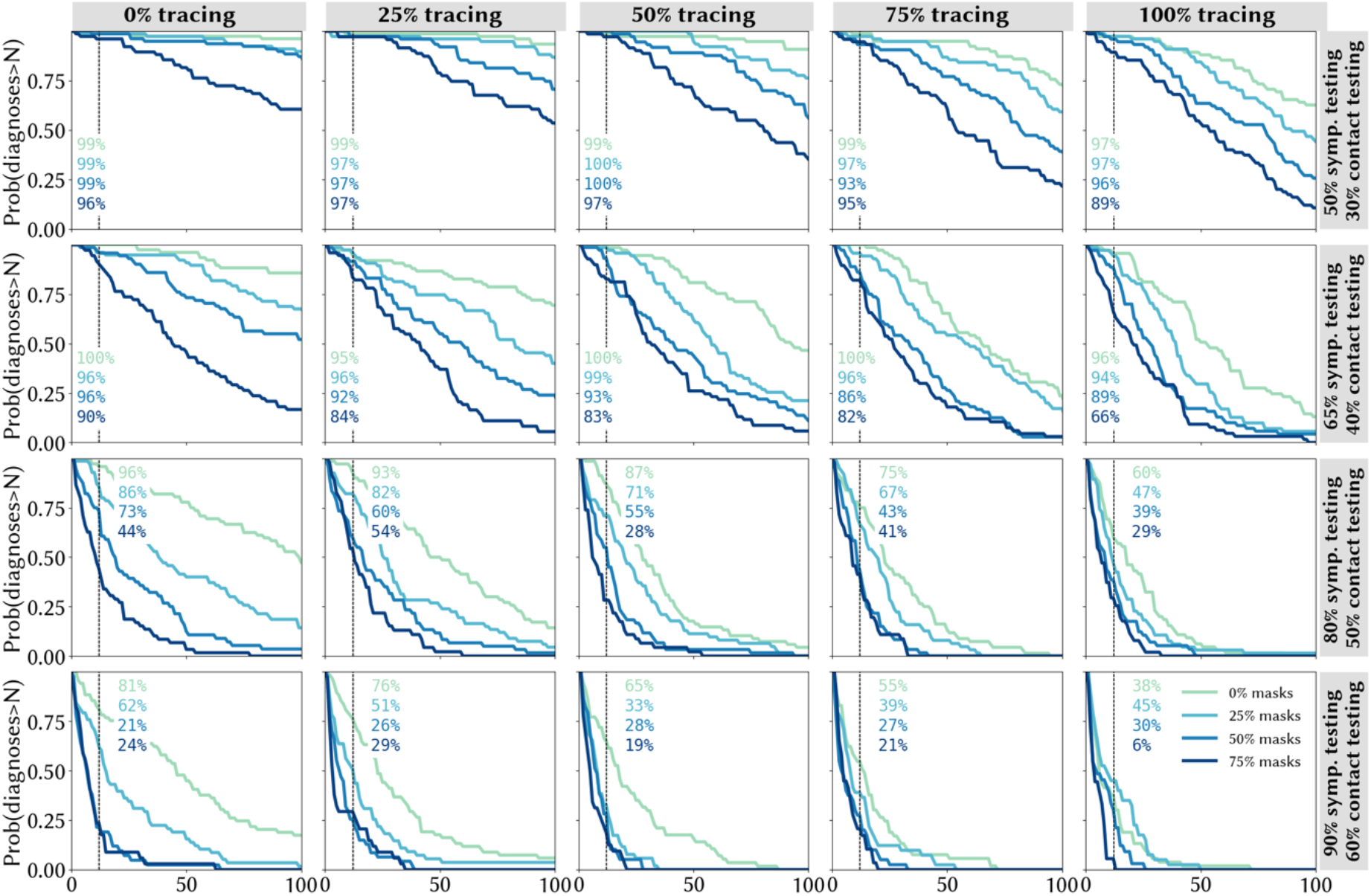
Sensitivity to asymptomatic testing assumption: the probability of the trailing 14-day average of locally-acquired cases exceeding a given number by December 31, 2020 with individual-level mask efficacy set to 15%, under different assumptions about the testing rate (rows), proportion of community contacts that can be traced within one week (columns), and mask uptake (line colours).

**Figure S3.**
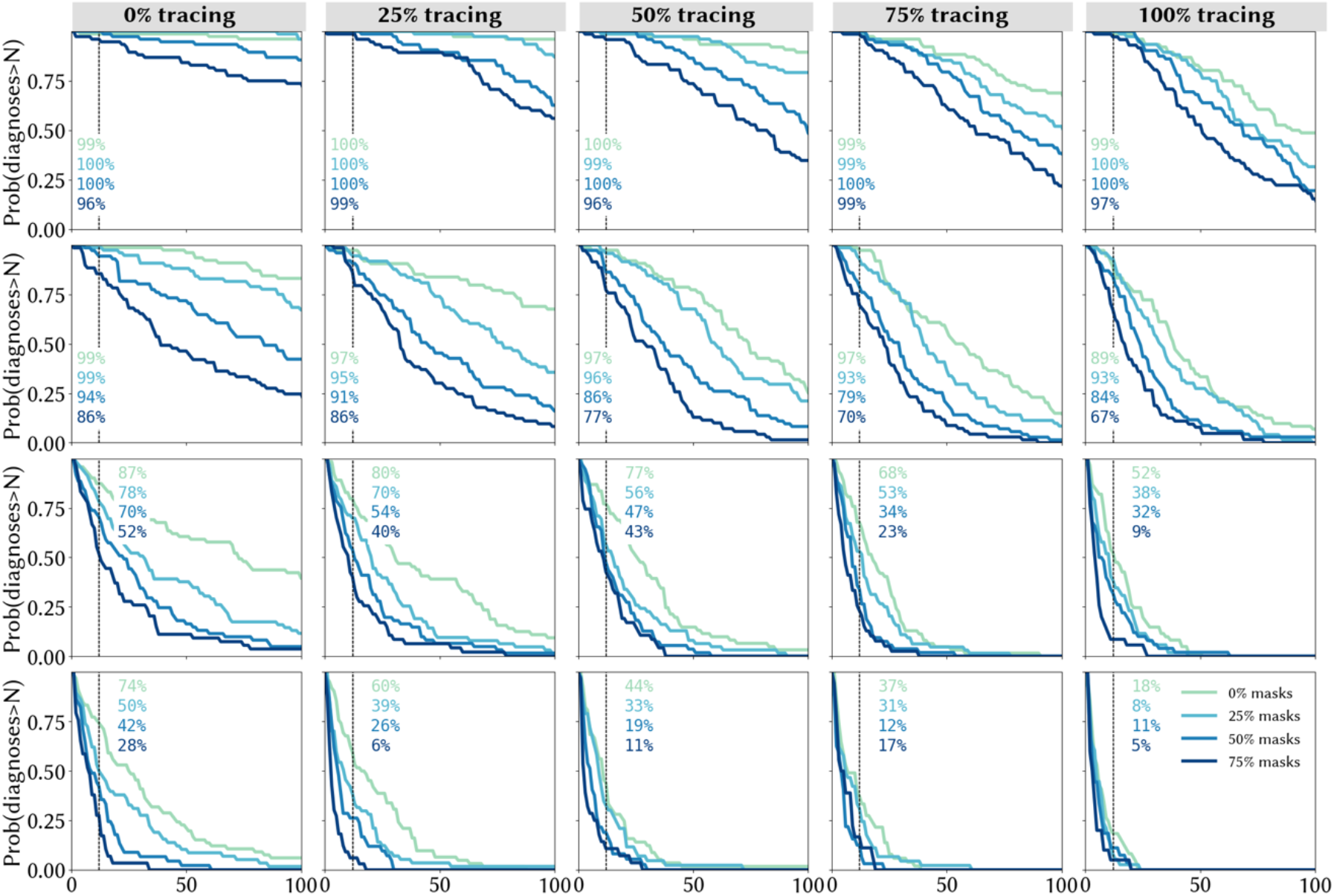
Sensitivity to mask efficacy assumption: the probability of the trailing 14-day average of locally-acquired cases exceeding a given number by December 31, 2020 with individual-level mask efficacy set to 15%, under different assumptions about the testing rate (rows), proportion of community contacts that can be traced within one week (columns), and mask uptake (line colours).

**Figure S4.**
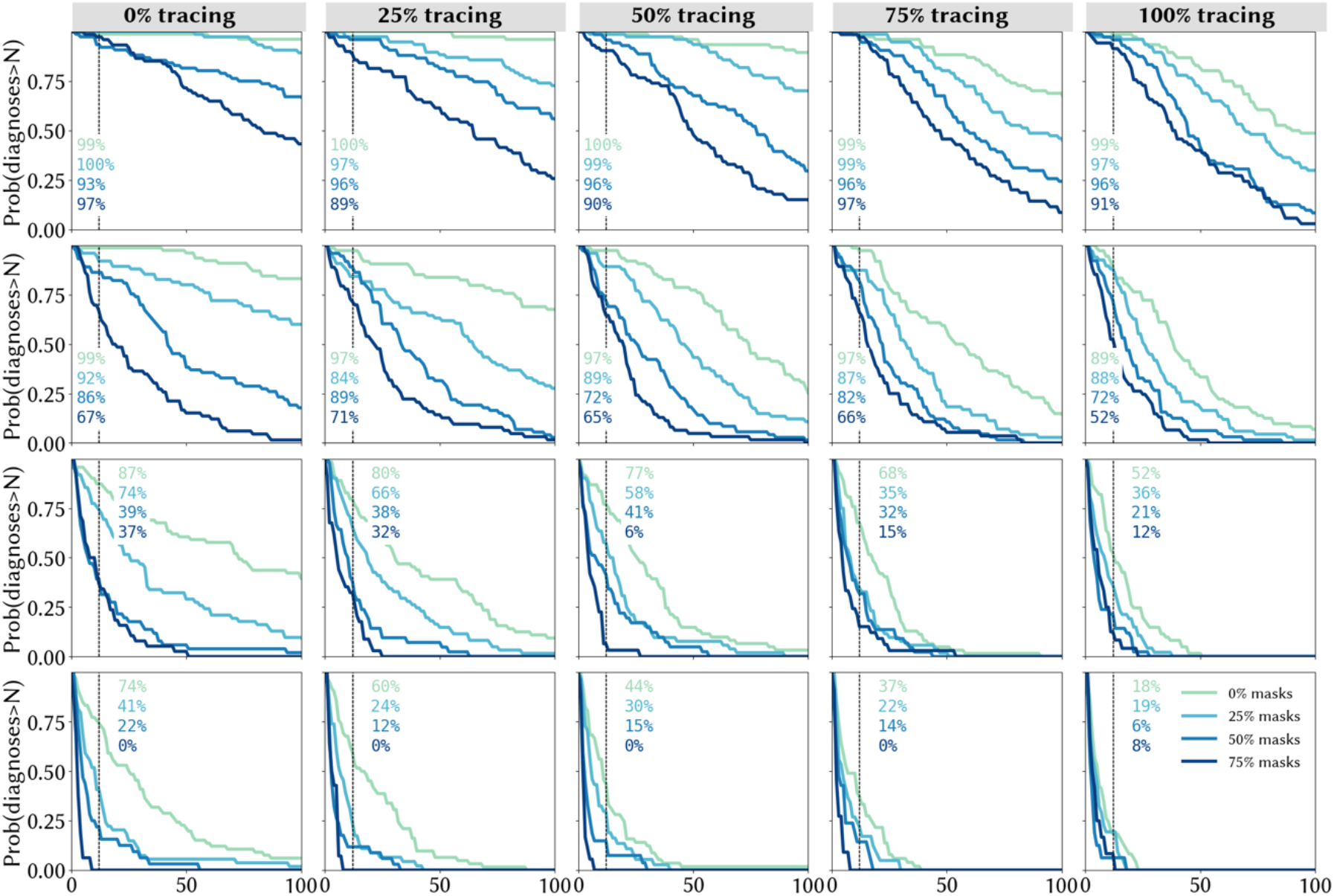
Sensitivity to mask efficacy assumption: the probability of the trailing 14-day average of locally-acquired cases exceeding a given number by December 31, 2020 with individual-level mask efficacy set to 45%, under different assumptions about the testing rate (rows), proportion of community contacts that can be traced within one week (columns), and mask uptake (line colours).

